# Mortality of COVID-19 is Associated with Cellular Immune Function Compared to Immune Function in Chinese Han Population

**DOI:** 10.1101/2020.03.08.20031229

**Authors:** Qiang Zeng, Yong-zhe Li, Gang Huang, Wei Wu, Sheng-yong Dong, Yang Xu

## Abstract

In December 2019, novel coronavirus (SARS-CoV-2) infected pneumonia occurred in Wuhan, China. The number of cases has increased rapidly but information on the clinical characteristics of SARS-CoV-2 pneumonia compared to normal controls in Chinese Han population is limited. Our objective is to describe the clinical characteristics of SARS-CoV-2 pneumonia compared to normal controls in the Chinese Han population. In this case series of 752 patients, the full spectrum of cases is described. Fever was present in 86-90% of the patients. The second most common symptom was cough (49.1-51.0%), fatigue (25.2-27.1%), sputum (20.0-23.1%), and headache (9.8-11.1%). the mortality rate is 4.6% in Wuhan, 1.9% in Beijing, and 0.9% in Shanghai. Our findings showed that the levels of lymphocytes were 0.8(IQR, 0.6-1.1)10^9^/L in Wuhan, 1.0(IQR, 0.7-1.4)10^9^/L in Beijing, and 1.1 (IQR, 0.8-1.5) 10^9^/L in Shanghai before admission to hospitals, respectively, indicating that cellular immune function might relate to the mortality. Based on the reference ranges of normal Chinese Han population and the data of the critically ill patients we have observed, it is recommended that reference ranges of people at high risk of COVID-19 infection are CD3^+^ lymphocytes below 900 cells/mm^3^, CD4^+^ lymphocytes below 500 cells/mm^3^, and CD8^+^ lymphocytes below 300 cells/mm^3^.

## Introduction

In December 2019, a cluster of acute respiratory illness, now known as SARS-CoV-2 pneumonia, occurred in Wuhan, China.^1-5^ The disease has rapidly spread from Wuhan to other areas. As of February 11, 2020, the Chinese Center for Disease Control and Prevention (China CDC) has officially reported that there are 2.0% (889) asymptomatic cases, 2.3% (1,023) death cases, and 80.9% mild cases among 44,672 confirmed cases. 51.4% (22,981) were male and 48.6% (21,691) were female.^6^

In January, 2020, the 2019 novel coronavirus (SARS-CoV-2) was identified in samples of bronchoalveolar lavage fluid from a patient in Wuhan and was confirmed as the cause of the SARS-CoV-2 pneumonia.^1,2^ Full-genome sequencing and phylogenic analysis indicated that SARS-CoV-2 is a distinct clade from the betacoronaviruses associated with human severe acute respiratory syndrome (SARS) and Middle East respiratory syndrome (MERS). ^1,2^

COVID-19 has spread rapidly since it was first identified in Wuhan and has been shown to have a wide spectrum of severity. Recently, a report shows that SARS-CoV and SARS-CoV-2 shared the same functional host-cell receptor, angiotensin-converting enzyme 2 (ACE2).^7^ Furthermore, SARS-CoV-2 binds to ACE2 receptors in 10-20 fold higher affinity than SARS-CoV binds to the same receptors.^7^ Another report shows SARS-CoV-2 cell entry depends on ACE2 and TMPRSS2.^8^ Since ACE2 receptors on lung alveolar epithelial cells and enterocytes of the small intestine are dominant, lung alveolar epithelial cells or enterocytes of the small intestine may be an important susceptibility factor for human.^9^

According to World Health Organization interim guidance on January 12, 2020, SARS-CoV-2 infection is classified as asymptomatic case, mild and severe cases of pneumonia, and critical cases of pneumonia (ARDS, sepsis, septic shock). Severe cases of pneumonia are defined as patients with respiratory rate > 30 breaths/min, severe respiratory distress, or SpO_2_ < 90% on room air.^10^ Asymptomatic case has been reported in China and Germany.^11,12^ Huang et al^3^ first reported 41 cases of SARS-CoV-2 pneumonia in which most patients had a history of exposure to Huanan Seafood Wholesale Market. Organ dysfunction (shock, acute respiratory distress syndrome [ARDS], acute cardiac injury, and acute kidney injury, etc.) and death can occur in severe or critical cases. Guan et al^4^ reported findings from 1099 cases of SARS-CoV-2 pneumonia and the results suggested that the SARS-CoV-2 infection clustered within groups of humans in close contact, and was more likely to affect older men with comorbidities. However, the full spectrum of disease without comorbidities or related to cellular immune functions is not yet known. The objective of this case series was to describe the clinical characteristics of SARS-CoV-2 pneumonia compared to 14,117 normal controls in Chinese Han population.

## Methods

### Study Design and Participants

This retrospective study was done at Chinese PLA General Hospital, Peking Union Medical College Hospital, and affiliated hospitals of Shanghai University of Medicine & Health Sciences. We extracted 752 case data in Shanghai (338 cases), and Beijing (414 cases) with clinical confirmed COVID-19 from China CDC. 752 case data were enrolled to be finally analyzed. Main symptoms and laboratory characteristics were from 178 patients surveyed by random sampling. This case series was approved by the institutional ethics board of Shanghai University of Medicine & Health Sciences and Peking Union Medical College Hospital (#ZS-1830). 14,117 normal controls from health checkup (from November 2018 to November 2019, before COVID-19 outbreak) in Chinese PLA General Hospital, Peking Union Medical College Hospital, and affiliated hospitals of Shanghai University of Medicine & Health Sciences were retrospectively enrolled. Identification of patients was achieved by reviewing and analyzing available electronic medical records and patient care resources. Written informed consent was waived due to the rapid emergence of this infectious disease. We retrospectively analyzed patients according to WHO interim guidance.^13^ Laboratory confirmation of SARS-CoV-2 infection was performed as previously described.^14^

All patients with SARS-CoV-2 pneumonia enrolled in this study were diagnosed according to World Health Organization interim guidance.^13^ According to World Health Organization interim guidance on January 12, 2020, SARS-CoV-2 infection is classified as asymptomatic cases, mild and severe cases of pneumonia, and critical cases of pneumonia (ARDS, sepsis, septic shock). Severe case of pneumonia is defined as patients with respiratory rate > 30 breaths/min, severe respiratory distress, or SpO_2_ < 90% on room air.^10^ Clinical outcomes (discharges, mortality, and recovery, etc.) were monitored up to March 4, 2020, the final date of follow-up.

### Real-Time Reverse Transcription Polymerase Chain Reaction Assay for SARS-CoV-2

A confirmed case of COVID-19 is defined as a positive result on high throughput sequencing or real-time reverse-transcriptase–polymerase-chain-reaction (RT-PCR) assay of pharyngeal swab specimens. Samples were collected for extracting SARS-CoV-2 RNA from patients suspected of having SARS-CoV-2 infection as described previously.^14^ In brief, the pharyngeal swabs were placed into a collection tube with 150 μL of virus preservation solution, and total RNA was extracted within 2 hours using the respiratory sample RNA isolation kit. Forty μL of cell lysates were transferred into a collection tube followed by vortex for 10 seconds. After standing at room temperature for 10 minutes, the collection tube was centrifugated at 1000 rpm/min for 5 minutes. The suspension was used for RT-PCR assay of SARS-CoV-2 RNA. Two target genes, including open reading frame 1ab (*ORF1ab*) and nucleocapsid protein (N), were simultaneously amplified and tested during the real-time RT-PCR assay. Target 1 (*ORF1ab*): forward primer CCCTGTGGGTTTTACACTTAA; reverse primer ACGATTGTGCATCAGCTGA; and the probe 5′-VIC-CCGTCTGCGGTATGTGGAAAGGTTATGG-BHQ1-3′. Target 2 (N): forward primer GGGGAACTTCTCCTGCTAGAAT; reverse primer CAGACATTTTGCTCTCAAGCTG; and the probe 5′-FAM-TTGCTGCTGCTTGACAGATT-TAMRA-3′. The real-time RT-PCR assay was performed using a SARS-CoV-2 nucleic acid detection kit. Reaction mixture contains 12 μL of reaction buffer, 4 μL of enzyme solution, 4 μL of probe primers solution, 3 μL of diethyl pyrocarbonate–treated water, and 2 μL of RNA template. RT-PCR assay was performed under the following conditions: incubation at 50 °C for 15 minutes and 95 °C for 5 minutes, 40 cycles of denaturation at 94 °C for 15 seconds, and extending and collecting fluorescence signal at 55 °C for 45 seconds. A cycle threshold value (Ct-value) less than 37 was defined as a positive test result, and a Ct-value of 40 or more was defined as a negative test. These diagnostic criteria were based on the recommendation by the National Institute for Viral Disease Control and Prevention (China). A medium load, defined as a Ct-value of 37 to less than 40, required confirmation by retesting. Only RT-PCR confirmed cases were included in the analysis.

### Flow Cytometry Data Analysis

All antibodies were obtained from BD Biosciences (San Jose, CA, USA). Two 100-μL samples of blood were placed in two tubes for staining according to mmanufacturer’s manual. After this procedure, 1 mL of red-cell lyses buffer was added to each tube and incubated for 10 minutes and then washed with Sorvall cell washer (Thermo Fisher Scientific, Waltham, MA, USA). Cells were then resuspended in 350 μL of phosphate-buffered saline and acquired with the use of FACSCanto*™* flow cytometry; daily quality control and assurance were carried out with the use of seven-color setup beads (BD Bioscience, San Jose, CA, USA).

### Statistical Analysis

Categorical variables were described as frequency rates and percentages, and continuous variables were described using mean, median, and interquartile range (IQR) values. Means for continuous variables were compared using independent group *t* tests when the data were normally distributed; otherwise, the Mann-Whitney test was used. Data (non-normal distribution) from repeated measures were compared using the generalized linear mixed model. Proportions for categorical variables were compared using the χ^2^ test, although the Fisher exact test was used when the data were limited. All statistical analyses were performed using SPSS (Statistical Package for the Social Sciences) version 13.0 software (SPSS Inc). For unadjusted comparisons, a 2-sided α of less than 0.05 was considered statistically significant. The analyses have not been adjusted for multiple comparisons and, given the potential for type I error, the findings should be interpreted as exploratory and descriptive. Because the cohort of patients in our study was not derived from random selection, all statistics are deemed to be descriptive only.

## Results

### Presenting Characteristics

The study population included 752 cases with confirmed SARS-CoV-2 pneumonia and 14,117 normal controls. Fever was present in 86-90% of the patients. The second most common symptom was cough (49.1-51.0%), fatigue (25.2-27.1%), sputum (20.0-23.1%), and headache (9.8-11.1%) (Table 1). Less common symptoms were chill, sore throat, shortness of breath, anorexia, diarrhea, nausea, and vomiting.

**Table 1.** Characteristics of COVID-19 patients in Shanghai and Beijing

| Contents | Shanghai<br>(n=338) | Beijing<br>(n=414) |
| --- | --- | --- |
| Main symptoms |  |  |
| Fever | 304 (90.0%) | 356 (86.0%) |
| Cough | 166 (49.1%) | 211 (51.0%) |
| Fatigue | 85 (25.2%) | 112 (27.1%) |
| Sputum | 78 (23.1%) | 83 (20.0%) |
| Sore throat | 54 (16.0%) | 70 (16.9%) |
| Headache | 33 (9.8%) | 46 (11.1%) |
| Main laboratory characteristics |  |  |
| White blood cell count, $10^9/L$ | 4.6 (3.6-5.9) | 4.6 (3.7-6.1) |
| Neutrophil count, $10^9/L$ | 2.9 (2.4-3.8) | 2.8 (1.2-3.4) |
| Lymphocyte count, $10^9/L$ | 1.1 (0.8-1.5) | 1.0 (0.7-1.4) |
| D-dimer, mg/L | 0.43 (0.28-0.85) | 0.5 (0.31-1.25) |
| C-reactive protein, mg/L | 9.8 (2.3-26.9) | 8.0 (1.8-25.1) |
| Interleukin-6, pg/mL | 11.0 (5.2-24.8) | 11.3 (5.5-25.1) |
| Prognosis |  |  |
| Death | 3 (0.9%) | 8 (1.9%) |
| Recovery | 298 (88.2%) | 294 (70.0%) |

The COVID-19 mortality rate in China is 2.3%.^6^ However, the mortality rate is 4.6% (2,282 death cases in 49,540 confirmed cases)in Wuhan, 1.9% (8 death cases in 414 confirmed cases) in Beijing, and 0.9% (3 death cases in 338 confirmed cases) in Shanghai based on China CDC official report on March 4, 2020. Our findings showed that the levels of lymphocytes were 0.8(IQR, 0.6-1.1)10^9^/L in Wuhan,^3^ 1.0(IQR, 0.7-1.4)10^9^/L in Beijing, and 1.1 (IQR, 0.8-1.5) 10^9^/L in Shanghai before admission to hospitals (Table 1), respectively, indicating that cellular immune function might relate to the mortality.

### Radiologic and Laboratory Parameters in Mild and Severe or Critical Patients

The radiologic and laboratory findings were showed (Table 1). These data could helpfully distinguish between mild and severe or critical patients. All of the 752 enrolled patients showed bilateral patchy patterns, or ground-glass opacity, or local patchy shadowing of chest CT scan. On admission, the predominant pattern of abnormality observed was bilateral patchy patterns (49.3%), local patchy shadowing (29.0%), and ground glass opacity (21.7%).

Due to lack of reference ranges for blood routine and lymphocytes of normal Chinese Han population, we systematically analyzed the 14,117 healthy adult Chinese Han people of 18-86 years old and classified them as an age group every 10 years for the convenience of other researchers in the future (Table 2).

**Table 2.** Characteristics of lymphocyte subsets in 14,117 normal Chinese Han population

| Contents | Units | <20<br>(n = 31) | 20-29<br>(n = 444) | 30-39<br>(n = 1766) | 40-49<br>(n = 5781) | 50-59<br>(n = 4801) | 60-69<br>(n = 1046) | 70-79<br>(n = 227) | >80<br>(n = 21) |
| --- | --- | --- | --- | --- | --- | --- | --- | --- | --- |
| CD3<sup>+</sup> | cells/mm<sup>3</sup> | 1835<br>(1370-1985) | 1648<br>(1375-1999) | 1502<br>(1233-1870) | 1417<br>(1161-1735) | 1379<br>(1123-1671) | 1309<br>(1064-1625) | 1220<br>(943-1549) | 938<br>(761-1021) |
| CD4<sup>+</sup> | cells/mm<sup>3</sup> | 857<br>(652-957) | 876<br>(714-1070) | 852<br>(680-1071) | 829<br>(663-1052) | 822<br>(647-1031) | 763<br>(606- 975) | 718<br>(542-964) | 530<br>(453- 662) |
| CD8<sup>+</sup> | cells/mm<sup>3</sup> | 621<br>(498-823) | 594<br>(470-790) | 526<br>(403-690) | 465<br>(359-599) | 428<br>(331-558) | 421<br>(305-575) | 391<br>(278-518) | 348<br>(308-390) |
| CD4<sup>+</sup>/CD8<sup>+</sup> | ratio | 1.24<br>(1.09-1.52) | 1.49<br>(1.19-1.84) | 1.64<br>(1.27-2.13) | 1.81<br>(1.39-2.33) | 1.92<br>(1.44-2.53) | 1.84<br>(1.35-2.50) | 1.82<br>(1.44-2.77) | 2.09<br>(0.94-2.47) |
| CD19<sup>+</sup> | cells/mm<sup>3</sup> | 304<br>(209-372) | 279<br>(201- 364) | 247<br>(176-346) | 238<br>(170-332) | 237<br>(170-322) | 230<br>(158-311) | 210<br>(124-290) | 166<br>(71- 269) |
| CD16<sup>+</sup>CD56<sup>+</sup> | cells/mm<sup>3</sup> | 257<br>(168-284) | 180<br>(118- 270) | 188<br>(131-273) | 189<br>(126-276) | 200<br>(137-294) | 244<br>(168-355) | 265<br>(180-356) | 304<br>(240-432) |

The reference ranges of lymphocyte subsets are as followings (Table 2): CD3^+^ lymphocytes (938-1835 cells/mm^3^), CD4^+^ lymphocytes (530-857 cells/mm^3^), CD8^+^ lymphocytes (348-621 cells/mm^3^), CD4/CD8 ratio (1.24-2.09), CD19^+^ lymphocytes (166-304 cells/mm^3^), and CD16^+^CD56^+^ lymphocytes (180-304 cells/mm^3^). As the age increases,the levels of CD4 lymphocytes, CD8 lymphocytes, and B lymphocytes were decreased, whereas, the levels of natural killer (NK) cells were elevated in Chinese Han population suggesting that the T and B cell immune functions decline with age (Table 2).

The phenomenon of lymphocyte depletion (PLD) observed in severe or critical cases (ICU) in 100% (Table 3). As the disease progressed and clinical status deteriorated, the levels of lymphocytes progressively decreased before death. Further analyzing 178 cases without glucocorticoid treatment, their lymphocyte subsets showed that CD4 and CD8 T lymphocytes have significant difference (*p* < 0.01) between mild (Non-ICU) and severe or critical (ICU) cases (Table 3).

**Table 3.**
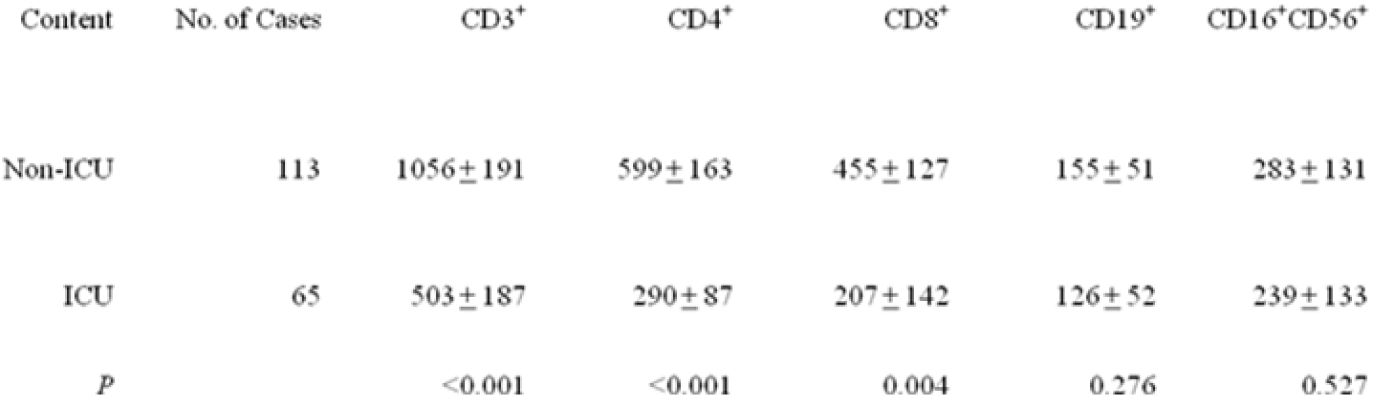
Lymphocytes of COVID-19 pneumonia in ICU and Non-ICU without glucocorticoid treatment

| Content | No. of Cases | CD3<sup>+</sup> | CD4<sup>+</sup> | CD8<sup>+</sup> | CD19<sup>+</sup> | CD16<sup>+</sup>CD56<sup>+</sup> |
| --- | --- | --- | --- | --- | --- | --- |
| Non-ICU | 113 | 1056 ± 191 | 599 ± 163 | 455 ± 127 | 155 ± 51 | 283 ± 131 |
| ICU | 65 | 503 ± 187 | 290 ± 87 | 207 ± 142 | 126 ± 52 | 239 ± 133 |
| *P* |  | <0.001 | <0.001 | 0.004 | 0.276 | 0.527 |

The levels of D-dimer, C-reactive protein and interleukin-6 were elevated (Table 1), indicating blood clot formation, severe inflammation, and cytokine storm in COVID-19 patients.

## Discussion

This report, to our knowledge, is the first study of COVID-19 with normal controls. The study population included 752 confirmed COVID-19 cases and 14,117 normal controls. Fever was present in 86-90% of the patients. The second most common symptom was cough (49.1-51.0%), fatigue (25.2-27.1%), sputum (20.0-23.1%), and headache (9.8-11.1%) (Table 1). Less common symptoms were chill, sore throat, shortness of breath, anorexia, diarrhea, nausea, and vomiting. The levels of D-dimer, C-reactive protein and interleukin-6 were elevated (Table 1), indicating blood clot formation, severe inflammation, and cytokine storm in COVID-19 patients.

The reference ranges of lymphocyte subsets are as followings (Table 2): CD3^+^ lymphocytes (938-1835 cells/mm^3^), CD4^+^ lymphocytes (530-857 cells/mm^3^), CD8^+^ lymphocytes (348-621 cells/mm^3^), CD4/CD8 ratio (1.24-2.09), CD19^+^ lymphocytes (166-304 cells/mm^3^), and CD16^+^CD56^+^ lymphocytes (180-304 cells/mm^3^). As the age increases,the levels of CD4 lymphocytes, CD8 lymphocytes, and B lymphocytes were decreased, whereas, the levels of natural killer (NK) cells were elevated in Chinese Han population suggesting that the T and B cell immune functions decline with age (Table 2). Based on the reference ranges of normal Chinese Han population and the data of the critically ill patients we have observed, it is recommended that reference ranges of people at high risk of COVID-19 infection are CD3^+^ lymphocytes below 900 cells/mm^3^, CD4^+^ lymphocytes below 500 cells/mm^3^, and CD8^+^ lymphocytes below 300 cells/mm^3^.

Secondary immunodeficiencies can result from various immunosuppressive agents (immunosuppressive drugs or glucocorticoids). Many specific diseases directly or indirectly cause secondary immunodeficiencies. This includes many types of cancer, and certain chronic virus infections (e.g. cytomegalovirus, varicella zoster virus). Our findings indicated the PLD at presentation were associated with the severity of disease (Table 3). The severe PLD or the secondary immunodeficiencies (CD4 lymphocytes are below 200 cells/mm^3^) is common in HIV patients. When CD4 lymphocytes are below 200 cells/mm^3^, patients with T cell depletion have increased susceptibility to fungal infections.^15,16^

It has been reported that SARS-CoV directly infects monocytes, macrophages and T lymphocytes in human.^17^ The PLD may also be secondary to activation of T lymphocytes.^18^ Our findings showed that CD4 and CD8 T lymphocytes have significant difference (*p* < 0.01) between mild (Non-ICU) and severe or critical (ICU) cases without glucocorticoid treatment indicating COVID-19 directly effected on immune system is possible.

COVID-19 has spread rapidly since it was first identified in Wuhan and has been shown to have a wide spectrum of severity as SARS-CoV-2 binds to ACE2 receptors in 10-20 fold higher affinity than SARS-CoV binds to the same receptors.^7,19^ The report shows SARS-CoV-2 cell entry depends on ACE2 and TMPRSS2.^8^ Therefore, it is necessary to further study whether lymphocytes have ACE2 receptor and TMPRSS2 expression during the developmental stage since the PLD exists. A SARS-CoV-2 RT-PCR assay for lymphocyte subsets in severe or critical cases should be followed up. Our findings also suggest that lymphocyte subsets should be analyzed at admission immediately no matter how COVID-19 affected lymphocytes through directly or indirectly. When patients are confirmed as COVID-19, clinicians should order tests for lymphocyte subsets in order to intervene early in the consequences of the PLD.

## Conclusions

In this case series of 752 patients, the full spectrum of cases is described. Fever was present in 86-90% of the patients. The second most common symptom was cough (49.1-51.0%), fatigue (25.2-27.1%), sputum (20.0-23.1%), and headache (9.8-11.1%). The mortality rate is 4.6% in Wuhan, 1.9% in Beijing, and 0.9% in Shanghai. Our findings showed that the levels of lymphocytes were 0.8(IQR, 0.6-1.1)10^9^/L in Wuhan, 1.0(IQR, 0.7-1.4)10^9^/L in Beijing, and 1.1 (IQR, 0.8-1.5) 10^9^/L in Shanghai before admission to hospitals, respectively, indicating that cellular immune function might relate to the mortality. Based on the reference ranges of normal Chinese Han population and the data of the critically ill patients we have observed, it is recommended that reference ranges of people at high risk of COVID-19 infection are CD3^+^ lymphocytes below 900 cells/mm^3^, CD4^+^ lymphocytes below 500 cells/mm^3^, and CD8^+^ lymphocytes below 300 cells/mm^3^.

## Data Availability

the availability of all data referred to in the manuscript is avalable

## Acknowledgment

We acknowledgment all volunteer emergency medical teams for Wuhan from Shanghai and Beijing.

